# Viral load dynamics in asymptomatic and symptomatic patients during Omicron BA.2 outbreak in Shanghai, China, 2022: a longitudinal cohort study

**DOI:** 10.1101/2024.01.23.24301654

**Authors:** Jingwen Ai, Jiaxin Zhou, Yang Li, Feng Sun, Shijia Ge, Haocheng Zhang, Yanpeng Wu, Yan Wang, Yilin Zhang, Hongyu Wang, Jianpeng Cai, Xian Zhou, Sen Wang, Rong Li, Zhen Feng, Xiangyanyu Xu, Xuemei Yan, Yuchen Zhao, Juanjuan Zhang, Hongjie Yu, Wenhong Zhang

## Abstract

**Backgrounds:** The SARS-CoV-2 virus has caused global outbreaks, including the recent Omicron BA.2 wave in Shanghai in spring 2022. While, the viral load dynamic of Omicron infections with different clinical severity was still unclear.

**Methods:** A prospective cohort was conducted on 48,830 hospitalized COVID-19 patients in three hospitals in Shanghai, China, from 23 March and 15 May 2022. Regular nucleic acid testing was performed and the Cycle threshold (Ct) value tested by RT-PCR was used as a proxy of viral load. The viral load dynamic by different clinical severity since initial detection and the risk factors were analyzed.

**Results:** The study included 31% asymptomatic cases, 68% mild-moderate cases, 1% severe cases, 1.29% critical and fatal cases. 57% of patients tested positive upon admission, average Ct value remained stable with peak viral concentrations occurring at 4 days (median Ct value of 27.5), followed by a decrease with a viral shedding time (VST) of 6.1 days (IQR, 4.0-8.8 days). Omicron viral load varied by age and clinical severity, but peak Ct values occurred at similar times. Unvaccinated status, age over 60, and comorbidities were associated with higher peak viral concentrations and longer shedding durations. Asymptomatic cases had a 40% contagious probability within 6 days of detection. Mild-moderate cases and severe cases had a 27%, >50% probability of infectiousness post-symptom resolution, respectively.

**Conclusion:** The initial Ct value could forecast severe consequences. Unvaccinated older people with specific comorbidities are the high-risk groups associated with high viral load and long shedding duration.

## Background

Coronavirus disease 2019 (COVID-19), caused by infection with severe acute respiratory syndrome coronavirus 2 (SARS-CoV-2), has resulted in unprecedented impact worldwide. Viral load, as a key determinant of viral shedding, is shown to correlate with infectivity, transmissibility, and pathogenicity. Understanding the SARS-CoV-2 viral load trajectory is crucial for developing epidemiological control strategies and optimizing clinical management of COVID-19 patients. The cycle threshold (Ct) value obtained by reverse transcription-polymerase chain reaction (RT-PCR) was widely used as a surrogate marker for infectiousness due to high correlation between RNA viral load and the presence of infectious virus^1^.

Since March 2022, a local outbreak of the Omicron sub-lineage BA.2^2, 3^ had occurred in Shanghai, resulting in over 0.6 million laboratory-confirmed infections of early June^4^. While previous studies have revealed some viral load characteristics of ancestral SARS-CoV-2 and other variants of concern (VOC) ^5–10^, Omicron variant has shown dramatic changes in viral dynamics and clinical features. Some previous studies mainly focused on the effect of vaccines on viral dynamics of Omicron or only analyzed in-depth the viral dynamics of symptomatic patients^11–15^. Several studies included asymptomatic individuals but the size numbers were limited^14, 16^ or only presented the viral shedding time^17^. One study analyzed the viral dynamics in detail focused on mild cases^15^. Evidences about peak viral load and viral shedding dynamics of Omicron are still sparse and there is a lack of multicenter evidence of viral dynamics on a broad spectrum of diseases including asymptomatic, mild and severe and death cases supporting by large sample sizes. This multicenter study aims to providing a comprehensive description of viral kinetic characteristics about Omicron BA.2 among patients with a wide disease spectrum ranging from asymptomatic to critically ill or death, and to identify the possible risk factors affecting viral characteristics.

## Methods

### Study design and participants

The study enrolled a total of 48830 COVID-19 cases, where all patients were admitted to the three designated hospitals/makeshift hospitals in Shanghai from 23 March to 15 May 2022, and their RT-PCR testing results were collected during the hospitalization. For each participant, serial samples of nasopharyngeal swabs were collected from all the participants during the hospitalization according to clinical need. Clinical information such as the vaccination history, including the vaccination dates and vaccine type, the date of illness onset, and duration of hospitalization were well documented from clinical records. Personal demographic information was also obtained from the questionnaire reported by patients or guardians.

COVID-19 cases were defined as individuals who tested positive for SARS-CoV-2 by RT-PCR. The degree of severity of the patient was categorized according to the clinical manifestation and the supportive measures used within the healthcare system according to a revised WHO clinical progression scale^18^. Specifically, the asymptomatic type had no clinical symptoms and no pneumonia manifestation can be found in imaging. The mild-moderate type had clinical symptoms or shows abnormalities in lung imaging and received no oxygen therapy. Patients who received oxygen therapy by mask or prongs, or received non-invasive ventilation (NIV) or high flow oxygen therapy would be categorized as severe type. Those who received intubation and mechanical ventilation or even died were defined as ‘Critical-death’ patients.

### Laboratory confirmation of COVID-19 by reverse-transcription polymerase chain reactions

The sampling and detection procedure was described in supplementary material. The (Ct) values of both ORF1ab and N genes of SARS-CoV-2 were collected but results are presented only for N gene since the patterns observed for both genes were similar (unpublished data). The Ct values of real-time RT PCR tests of nasopharyngeal species were used as surrogates of relative viral genomic loads, and Ct ≤35 was considered positive. The viral load level was inversely correlated with Ct value which is a low viral load characterized by a high Ct and a high viral load characterized by low Ct values. Additionally, several researches revealed that the cultural positive proportion of omicron virus from respiratory tract specimens was under 5% with Ct value of 32 ^16, 19–23^, proving the unsuccessful to recover infectious virus when Ct value of 35. Thus, we chose the Ct value of 35 as positive threshold to exploring the correlation between CT value and infectiveness in our study.

### Statistical analysis

Demographic information including age, gender, status of underlying disease, vaccination history at admission of patients was described in total as well as by clinical severity. Baseline characteristics among four clinical severity groups was compared using Chi-square test for discrete variables and the Wilcoxon test for continuous variables.

The kinetic of viral load of patients since initial detection was described by total, by clinical severity, by age groups, by vaccination status, and by comorbidities using mean and 95% confidence interval (CI). Virus shedding time (VST) was defined as the interval between the date of initial detection of PCR and the date of subsequent first negative PCR test. For patients with a positive result at their last follow-up, VST was right censor and was defined as the interval between the date of the first positive PCR test and date of the last positive test. The intervals of VST, intervals between symptom onset and first PCR positive, and the intervals between symptom resolution and first PCR positive were estimated using gamma, Weibull, and lognormal distributions based on AIC considering censor data. The median and interquartile range (IQR) were described for each interval.

The viral proliferation stage and viral clearance stage were classificated by the peak concentration of viral load, with the former is the phase before peak and the latter is the phase after peak. Thus, peak concentration of viral load and VST were described as essential index of the kinetic by total and by several factors mentioned above respectively. Multivariate linear regression and interval regression were used to investigate the risk factors associated with peak Ct value and VST. The patients with long VST (> 30 days) were excluded in baseline analysis. The comparison of baseline characteristics between participants and excluded patients was conducted. The statistical analyses were conducted by R (version 4.14).

## Results

As of May 15^th^, 2022, 48830 participants were included, with 31% of asymptomatic participants, 68% of mild-moderate cases, 1% severe cases, 0.29% of critical cases or death cases, respectively. The baseline characteristics of participants at admission were described by clinical status in table 1. The median age was 45 years old (IQR, 32-55 years) and 63% of participants were female. 21% of patients had at least one comorbidity, with hypertension (14%), diabetes (5%) and cardiovascular diseases (2%) being the top three comorbidities. The majority (88%) of patients had at least one-dose vaccination. Patients with severity of severe or above were older and with more comorbidities, but less vaccination coverage than asymptomatic and mild-moderate cases (*p*<0.001). The excluded patients had worsened clinical severity status but similar gender proportion and age distribution comparing to the participants (table S1).

**Table 1.** Baseline characteristics of enrolled participants by clinical severity.

| Characteristics | Total<br>(n=48830) | Asymptomatic<br>(n=15165,<br>31%) | Mild-moderate<br>(n=33038, 68%) | Severe<br>(n=487, 1%) | Critical or death<br>(n=140, 0.29%) | P value |
| --- | --- | --- | --- | --- | --- | --- |
| **Gender** |  |  |  |  |  | <0.001 |
| Female | 30664 (63) | 10081 (66) | 20220 (61) | 271 (56) | 92 (66) |  |
| Male | 18166 (37) | 5084 (34) | 12818 (39) | 216 (44) | 48 (34) |  |
| **Age (years)** |  |  |  |  |  | <0.001 |
| Median (IQR) | 45 (32-55) | 47 (34-56) | 43 (32-54) | 73 (65-85) | 78 (69-89) |  |
| 0-17 | 1811 (4) | 565 (4) | 1245 (4) | 1 (0) | 0 (0) |  |
| 18-59 | 39836 (82) | 11975 (79) | 27779 (84) | 66 (14) | 16 (11) |  |
| 60-79 | 6536 (13) | 2480 (16) | 3750 (11) | 249 (51) | 57 (41) |  |
| 80+ | 647 (1) | 145 (1) | 264 (1) | 171 (35) | 67 (48) |  |
| **Comorbidities** |  |  |  |  |  | <0.001 |
| None | 38093 (78) | 12166 (80) | 25871 (78) | 51 (10) | 5 (4) |  |
| Hypertension | 6872 (14) | 1973 (13) | 4481 (14) | 317 (65) | 101 (72) |  |
| Diabetes | 2336 (5) | 708 (5) | 1397 (4) | 164 (34) | 67 (48) |  |
| Cardiovascular disease | 1054 (2) | 227 (1) | 642 (2) | 117 (24) | 68 (49) |  |
| Hematologic diseases | 80 (0) | 11 (0) | 55 (0) | 10 (2) | 4 (3) |  |
| Kidney disease (Level1) | 172 (0) | 36 (0) | 108 (0) | 16 (3) | 12 (9) |  |
| Kidney dialysis (Level2) | 215 (0) | 29 (0) | 77 (0) | 80 (16) | 29 (21) |  |
| Kidney transplantation (Level3) | 13 (0) | 1 (0) | 6 (0) | 5 (1) | 1 (1) |  |
|  |  |  |  |  | 48 (34) |  |
| Neurological conditions | 412 (1) | 99 (1) | 187 (1) | 78 (16) |  |  |
| Pneumonia | 522 (1) | 91 (1) | 390 (1) | 38 (8) | 3 (2) |  |
|  |  |  |  |  | 0 (0) |  |
| HIV | 3 (0) | 1 (0) | 2 (0) | 0 (0) |  |  |
|  |  |  |  |  | 21 (15) |  |
| Malignancy | 345 (1) | 54 (0) | 227 (1) | 43 (9) |  |  |
|  |  |  |  |  | 8 (6) |  |
| Liver disease | 330 (1) | 58 (0) | 258 (1) | 6 (1) |  |  |
|  |  |  |  |  | 1 (1) |  |
| Rheumatism | 184 (0) | 30 (0) | 143 (0) | 10 (2) |  |  |
|  |  |  |  |  | 0 (0) |  |
| Rhinitis | 173 (0) | 17 (0) | 156 (0) | 0 (0) |  |  |
|  |  |  |  |  | 5 (4) |  |
| Psychotic condition | 86 (0) | 23 (0) | 56 (0) | 2 (0) |  |  |
| Smoking history | 5096 (10) | 1236 (8) | 3827 (12) | 31 (6) | 2 (1) |  |
| Other underlying disease | 6524 (13) | 1336 (9) | 4738 (14) | 332 (68) | 133 (95) |  |
| Missing | 574 (1) | 289 (2) | 269 (1) | 14 (3) | 2 (1) |  |
| **Vaccination type** |  |  |  |  |  | <0.001 |
| Unvaccinated | 5502 (11) | 1823 (12) | 3273 (10) | 328 (67) | 78 (56) |  |
| Inactivated_Partial (1 dose) | 1483 (3) | 503 (3) | 968 (3) | 10 (2) | 2 (1) |  |
| Inactivated_Full (2 doses) | 16848 (35) | 5273 (35) | 11516 (35) | 54 (11) | 5 (4) |  |
| Inactivated_booster (3 doses) | 22694 (46) | 6804 (45) | 15838 (48) | 39 (8) | 13 (9) |  |
| Ad_Full (1 dose) | 937 (2) | 335 (2) | 602 (2) | 0 (0) | 0 (0) |  |
| Ad_booster (2 doses) | 442 (1) | 153 (1) | 287 (1) | 1 (0) | 1 (1) |  |
| Protein_Partial (<3 doses) | 33 (0) | 5 (0) | 28 (0) | 0 (0) | 0 (0) |  |
| Protein_Full (3 doses) | 292 (1) | 42 (0) | 248 (1) | 2 (0) | 0 (0) |  |
| Mixed_Partial/Full (2/3 doses) | 12 (0) | 3 (0) | 9 (0) | 0 (0) | 0 (0) |  |
| Mixed_Booster (3 doses) | 13 (0) | 4 (0) | 9 (0) | 0 (0) | 0 (0) |  |
| Missing | 574 (1) | 220 (1) | 260 (1) | 53 (11) | 41 (29) |  |
| **Days since full vaccination** |  |  |  |  |  |  |
| (median, IQR) | 320 (291, 346) | 321 (294, 346) | 319 (289, 346) | 320 (298, 349) | 328 (310,342) | <0.001 |
| **Days since boost vaccination** |  |  |  |  |  |  |
| (median, IQR) | 113 (92, 137) | 113 (93, 135) | 112 (92, 137) | 116 (93, 131) | 119 (98, 133) | <0.001 |

383 All variables are expressed as n (%) except when indicated otherwise. IQR: interquartile range.

### Viral shedding dynamic and viral load concentration

On the day of initial detection 57% of patients were positive with an average Ct value of 33 (95% CI, 31-34). This data remained stable within the first 4 days, before the viral load declined significantly on an average of 6.1 days (IQR, 4.0-8.8 days), and slowly decreased further within the next 3 weeks (Fig. 1A, B). The corresponding negative proportion would also gently approach 100% after a rapid growth to 74% (95% CI, 73-74) by day 10 (Fig. 1A). The peak occurrence of viral concentrations was within the first 4 days (IQR, 3-6 days) with a median Ct value of 27.5 (IQR, 23.4-32.1) (Fig. 1C).

**Fig 1.**
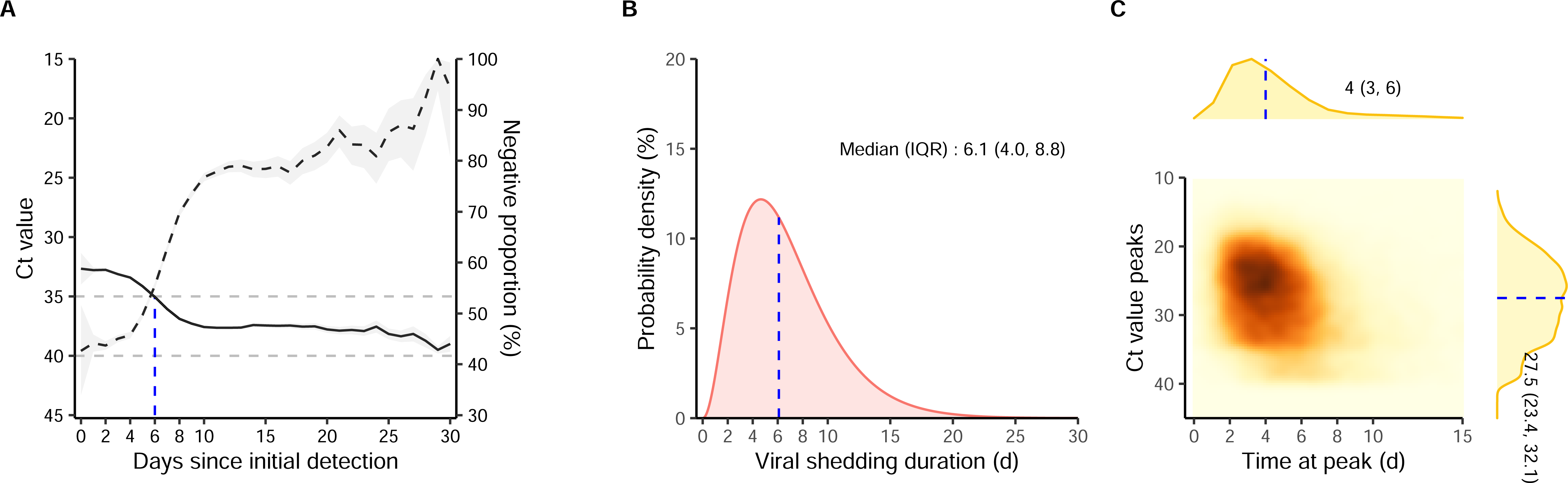
Characteristics of viral shedding of total participants. (A) Ct value and negative proportion at each time point since initial detection. Solid and dashed lines indicate average Ct value and proportion, respectively. Grey shadow indicates 95%CI. (B) Time distribution of viral shedding duration. Vertical dashed line indicates median of viral shedding time. (C) Correlation between peak Ct value and the time to reach peak since initial detection. Blue dashed lines indicate median of time at peak and the median of peak Ct value.

### Severity-specific viral shedding dynamic

Patients with worsened clinical severity were observed to have higher viral loads with average Ct values of 35 (95% CI, 33-37), 32 (95% CI, 27-40), 27 (95% CI, 20-34) for asymptomatic cases, mild-moderate, and severe patients, respectively, on the initial detection day, and 21 (95% CI, 20-27) for critical patients one day since initial detection (p < 0.001). As the virus was cleared, this superiority decreased with similar Ct values among the four groups after 24 days, except for similar Ct values between severe and critical groups during 0-5 days (Fig. 2A). The viral load of patients in the critical group showed a monotonically diminishing from a Ct value of 21 to 38 (95% CI, 37-40) with the longest viral shedding duration of 14.5 days (IQR, 7.9-23.4) compared to other groups. Conversely, for asymptomatic, mild-moderate, and severe cases, the average viral load was stable or significantly increasing within 4, 2, and 4 days, respectively, then dropped significantly with corresponding Ct values turning negative by day 5.2 (IQR: 3.2-8.0), 6.4 (IQR: 4.4-9.0), and 12.7 (IQR: 8.6-17.1). For patients with positive Ct values, the times of peak Ct value occurrence were similar among different groups (4-5 days), but peak concentrations showed gradual differences among the four clinical severity groups (p<0.001) (Fig. 2C).

**Fig 2.**
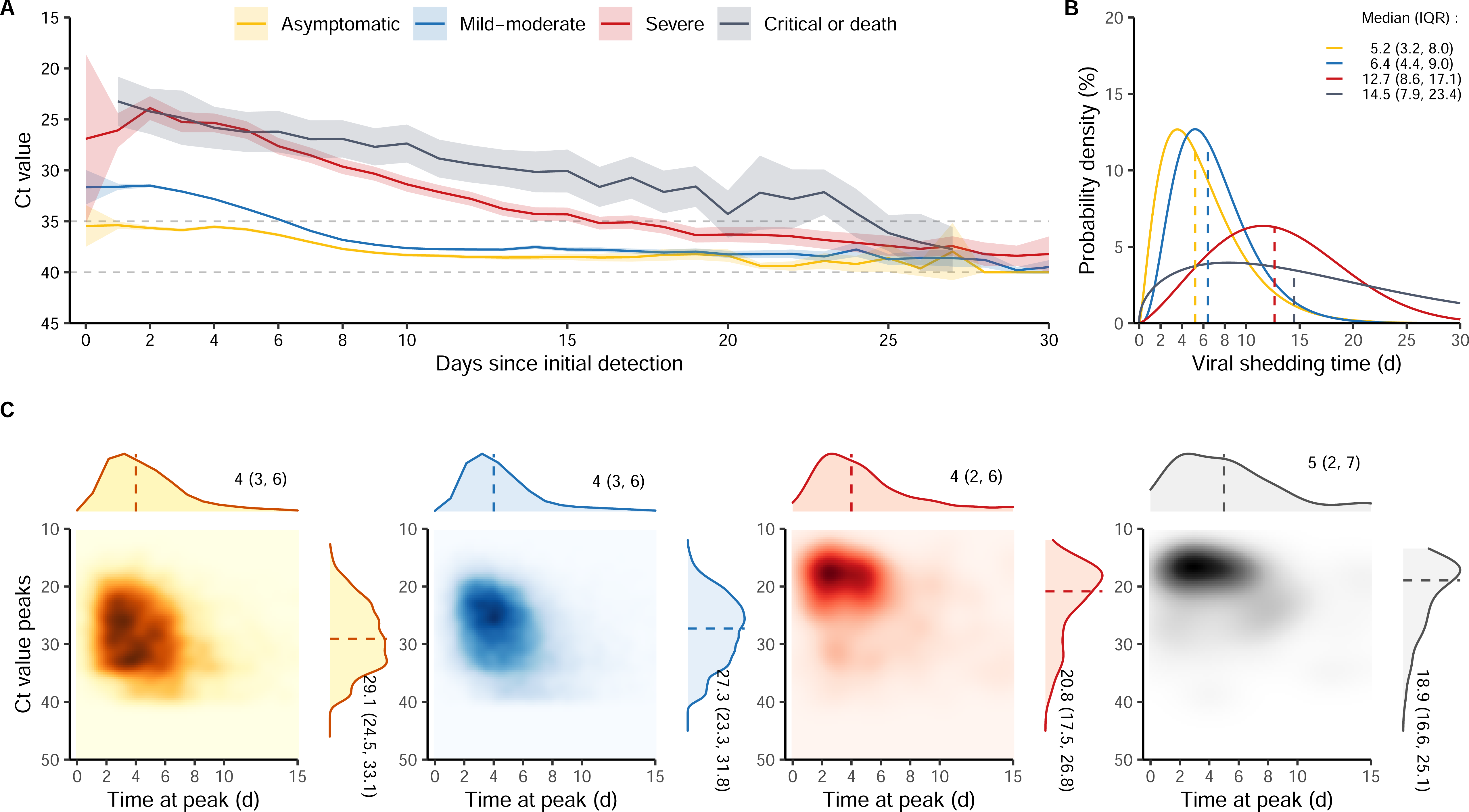
Clinical severity-specific characteristics of viral shedding. (A) Ct value at each time point since initial detection. Solid line indicates average Ct value, shadow indicates 95% CI. (B) Time distribution of viral shedding duration. Vertical dashed line indicates median of viral shedding time. (C) Correlation between peak Ct value and the time to reach peak since first positive PCR test. Vertical dashed line indicates median of time at peak and the median of peak Ct value.

**Fig 3.**
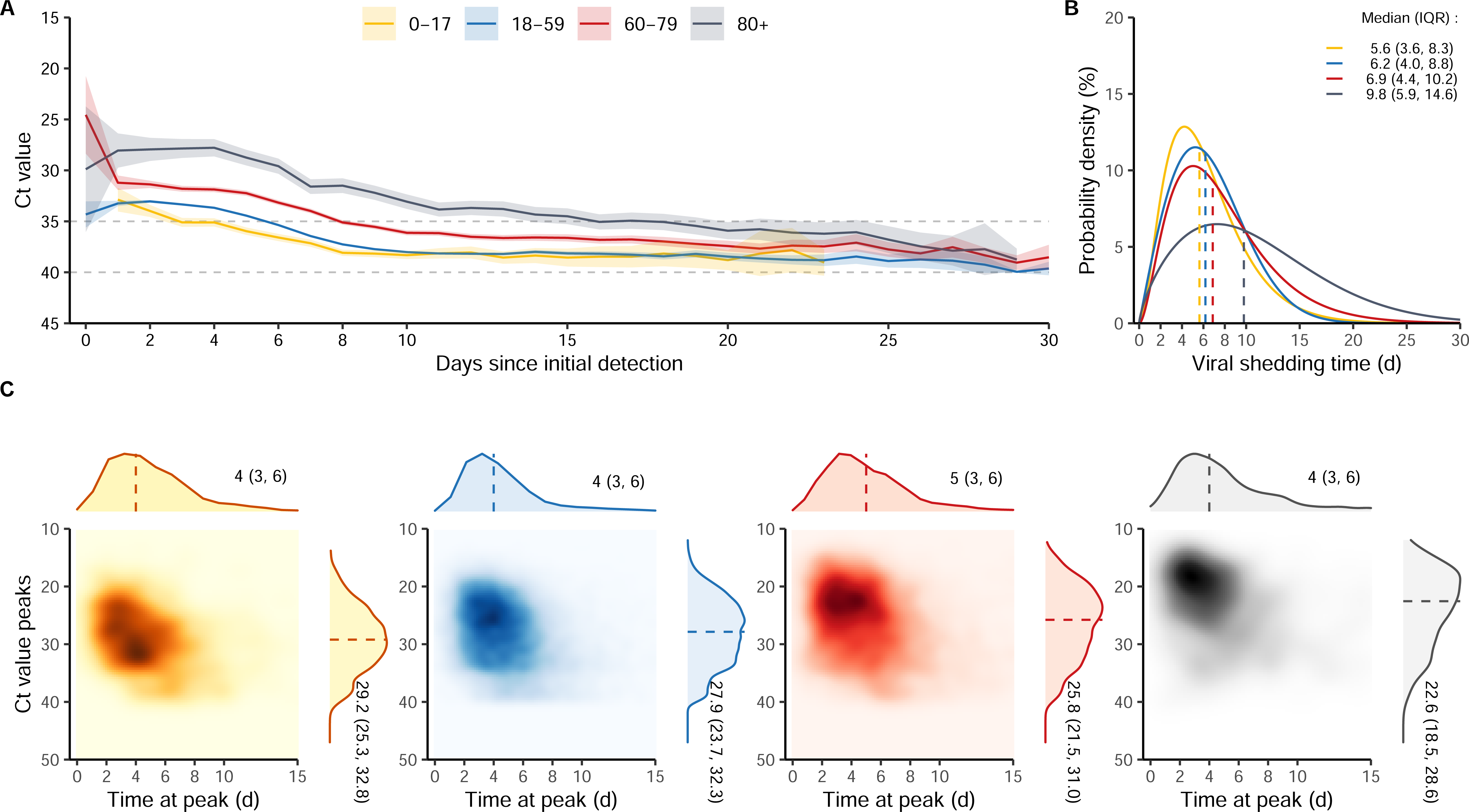
Age group-specific characteristics of viral shedding. (A) Ct value at each time point since first positive PCR test. Solid line indicates average Ct value, shadow indicates 95% CI. (B) Time distribution of viral shedding duration. Vertical dashed line indicates median of viral shedding time. (C) Correlation between peak Ct value and the time to reach peak since first positive PCR test. Vertical dashed line indicates median of time at peak and the median of peak Ct value.

### Age-specific viral shedding dynamic

Age-specific viral shedding dynamics revealed that elder patients had higher viral load and a longer viral shedding duration than the youngesters during most days of viral shedding, despite the lack of significant grade trend for four age groups in the first days (Fig. 2). On the initial detection day, the Ct value of the 60-79 years old group was lower than that of the 80+ years old group (25, 95% CI: 21-28; 30, 95% CI: 24-36, p=0.185), and was similar between the other age groups (33, 95% CI: 32-34 on day 1; 34, 95% CI: 34-36 on day 0, p = 0.366). Patients aged 80+ years old had the highest viral load, which increased within the first 4 days followed by a gentle decrease before conversion by day 9.8 (IQR, 5.9-14.6). Patients aged 18-59 and 60-79 years old experienced a VST of 6.2 days (IQR, 4.0-8.8) and 6.9 days (IQR, 4.4-10.2), respectively. In contrast, patients under 18 years old had a shortest shedding duration of 5.6 days (IQR, 3.6-8.3). Moreover, for patients with positive CT values, the time of peak CT value occurrence was similar among different groups (4-5 days), but peak viral concentrations showed an increasing trend with age (p<0.001). The results of viral shedding dynamic stratified by age and clinical severity were described in supplementary material.

### Factors associated with reduction in peak viral load and durations of viral shedding time

According to the multivariate analysis, higher peak viral concentrations and longer durations of effective viral shedding were observed in unvaccinated cases, those of older age, and with comorbidities (Fig. 4). Hypertension, kidney disease (especially kidney dialysis and kidney transplantation), neurological conditions, rheumatism and psychotic conditions were significantly correlated with higher peak viral load concentrations. Receiving at least one-dose vaccination was significantly associated with a reduction in peak viral load and a shorter viral shedding duration compared to the unvaccinated group. Sequential booster immunization was found to be more effective in reducing peak viral load and shortening viral shedding duration than homologous booster immunization, regardless of the type of vaccine (inactivated, ad-vectored, or protein subunit). Stratified analyses by clinical severity were reported in the supplementary material.

**Fig 4.**
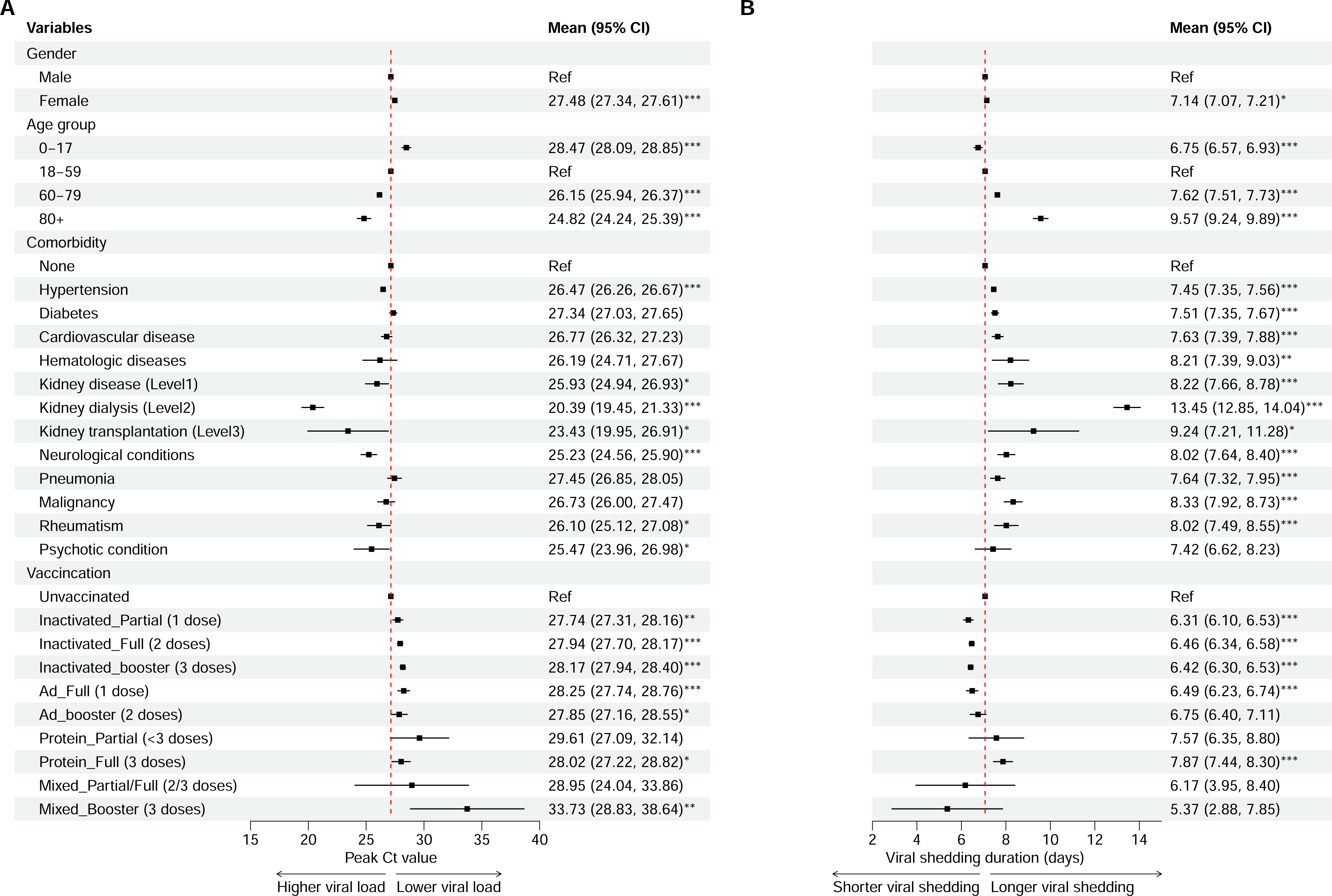
Risk factors associated with peak viral load (A) and viral shedding duration (B). The coefficients (dots) and 95% confidence intervals (95% CIs, line segments) were estimated from a multivariable regression analysis with an adjustment for gender, age, and comorbidities, and vaccination-infection status. Regression coefficients along with 95%CIs are reported as solid dots and horizontal lines relative to the value of the regression intercept. * p<0.05; ** p<0.01; *** p < 0.001.

### Relationship between infectivity and symptomatic status

Figure 5 illustrated the clinical severity-specific kinetic of positive proportion among patients since initial detection. Asymptomatic cases had a positive proportion of 34-39% within 6 days since initial detection, which decayed to 12% (95%CI: 10-15) after two weeks. For symptomatic cases, virus had already proliferation before symptom onset with lead times of 2.99, 1.56, and 2.13 days, resulting in high positive proportion of 63-66%, 67-85%, and 100% before onset for mild-moderate, severe, and critical or death patients, respectively. For mild-moderate and severe patients, the proportion decayed significantly as symptom subsided. Mild-moderate patients decreased to 27% (95%CI: 26-28) on the day of symptom resolution and below 20% after 14 days, while severe cases decayed to 76% (95%CI:70-80) on the day of symptom resolution and below 20% after 24 days. In the critical or death group, positive proportion maintained above 70% even after symptom resolution, until 20-30 days when it decreased to 46% (95%CI: 37-55).

**Fig 5.**
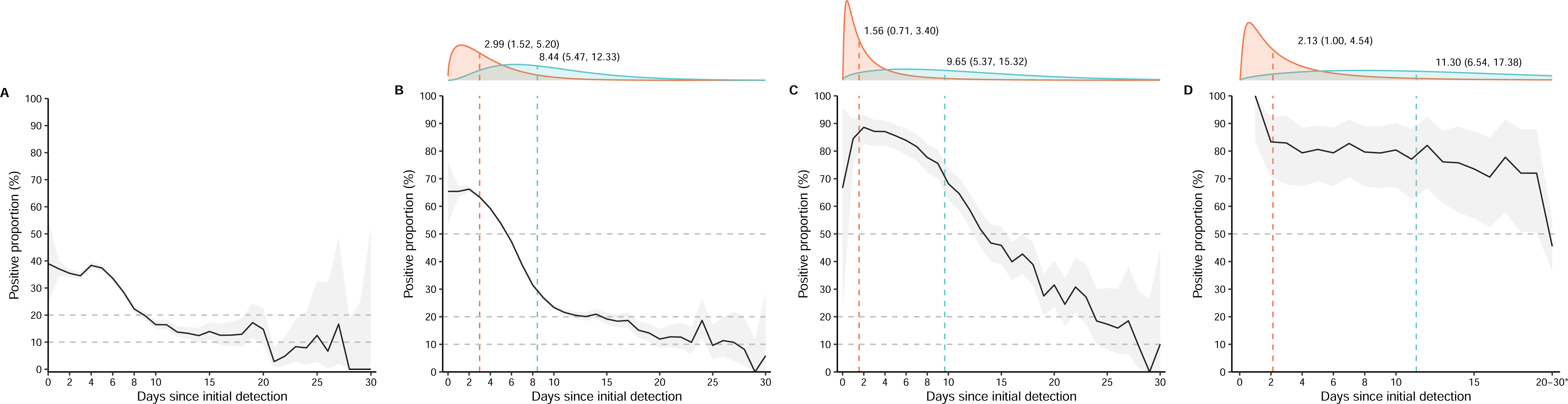
Clinical severity-specific progression of positive proportion from initial detection along with symptom statues. A: asymptomatic group; B: mild-moderate group; C: severe group; D: clinical or death group. The above margined figure is the density of time intervals between symptom onset since initial PCR detection (red) and the symptom resolution since initial PCR detection (blue). The vertical dashed lines represented the median of time of onset and symptom resolution date since initial detection. *: aggregated 20-30 days into a group, other time points were not treated.

## Discussion

Here we provided a comprehensive description of the characteristics of Omicron BA.2 viral shedding across a wide range of disease spectra and age groups, using Ct value as a surrogate of viral load. Among 48830 participants, 57% were PCR test positive on the initial detection day, and viral load then increased significantly to peak at around 4 days after detection following a decreasing trend with a VST of 6.1 days. Clinical severity and age had a clear influence on the viral shedding dynamics, as evidenced by the initial Ct value, peak Ct value and viral shedding duration, although the peak times were similar at 4-5 days.

It’s important to note that the study participants were from the general population and couldn’t be representative of immunocompromised individuals. Additionally, the viral load dynamic for the overall study population was relatively low and shedding quickly due to the large proportion of asymptomatic and mild-moderate cases with lower viral loads. Compared to the limited results on Omicron, our findings regarding the initial Ct value were similar to observations of adolescent children^24, 25^, adults^26^ and elderly patients^27^. Few studies have reported the severity-specific Omicron viral load, and the participants in these studies had different compositions of clinical severity^11, 15^. Lin’ study focus on the effect of vaccines and it’s hard to compare their results with this study^11^. The initial Ct value of asymptomatic and mild cases was higher than Yang’ s study, which may be explain by the earlier isolation in our study. The initial Ct value of asymptomatic cases was similar to Wu’s study, while the Ct value of symptomatic patients was relatively higher than that, which may be due to the high consistence of fatal cases in their study and the 2-day delay in specimen collection since onset ^28^. The peak viral load of mild-moderate patients in our study was similar to Hay’s study ^15, 29^. The average peak times reported were 2.5-4.2 days since detection and 3-4 days since onset ^15, 21, 30, 31^, and the viral load turned negative after 6-8 days, which could be prolonged to more than 10 days with older ages ^17, 28, 32^. These results were similar to our findings. In addition, the Ct value both at initial and the peak viral load stage in our study were higher than those of ancestral, Alpha, and Delta variant in the same population^28, 29, 33, 34^. This phenomenon has been frequently observed ^1, 19, 20^, indicating that high transmissibility of Omicron may be attributed to other mechanisms beyond viral load.

Accordingly, the overall Omicron infected individuals exhibited high viral load within the first week since initial detection indicating the highest infectious ability in the first week, a 14-day quarantine for infected people was an effective control as prior conducted^35^. The probability of infectious among asymptomatic cases was lowest during shortest infection duration comparing to symptomatic cases. For mild-moderate cases, the infectious probability had decayed to low level (20%) after symptom resolution. In the contrast, severe or above patients had extreme high infectious proportion during more than 24 days infectious durations and more than half of severe or above patients were still infectious after symptom resolution. Despite the relaxation of national COVID-19 control measures in many countries, certain institutions and workplaces, such as aging healthcare facilities, schools, and border entry points, require ongoing vigilance with respect to the virus. For these settings, we would recommend implementing graded control measures that take into account the clinical severity and updating the duration of isolation or self-monitoring accordingly to reduce the impact on normal life.

According to our results, patients with worsened clinical outcomes featured a lower initial Ct value post detection, peak value as well as longer shedding duration. Older age group, unvaccination, with comorbidities including hypertension, kidney disease especially kidney dialysis and kidney transplantation, neurological disorders, rheumatism, psychotic conditions and females were both independent risk factors associated with higher peak viral concentration as well as longer shedding duration, which were supported by previous studies^21, 36–39^. A delay or dysfunction in the initial triggering of the immune reaction led by aging ^40^, and higher levels of angiotensin converting enzyme 2 in alveoli of older people^41^, which is believed to be a receptor for novel coronaviruses, can partially explain the phenomenon. Our results support that getting at least one dose of vaccination and getting mixed booster could effectively reduce the peak viral load and shorten the shedding duration than unvaccinated patients.

Virus had proliferation preceding symptom onset and exhibited fastest replication in the first 4-5 days since detection, implying the requirement of antiviral therapy in early phase. Previous study indicated that the effect of nirmatrelvir/ritonavir treatment had minor clinical effect if practiced later than 5 days after onset^42^, and Paxlovid lacked efficacy for severe adult patients in preventing illness progressing^43^. Considering the delay of onset and detection, according our study, it is recommended giving timely antiviral drug within 3 days since onset for mild-moderate patients, which may play better effect on decrease peak level to shorten shedding duration and deduce risk developing severe outcome, particularly for high-risk group.

This study is the most comprehensive dynamic description about the viral shedding characters of omicron in Shanghai outbreak, 2022 covering all age groups and wide disease spectra with a sample size of over 40,000. In this study, we provided a comprehensive depiction of viral shedding pattern stratified by clinical severity, age groups, number of comorbidities, and vaccination status, and a quantified analysis to investigate the independent risk factors associated viral shedding characters and estimation the effect. However, there are still some weaknesses in our study. Firstly, this is a site-selection study, although the large-sample size, the participants could not represent the entire population in Shanghai, China. Secondly, the clinical diagnosis information on admission was incompletely collected making it impossible to respond the relationship between symptom progression and viral load dynamic in individual level. Thirdly, due to unavoidable reasons, the outcome of viral conversion for critical or death groups was more likely censored, resulting in fluctuation in Ct dynamic in the later phase of onset and wide confidence interval of VST estimation.

This study found that using Ct value as the proxy viral load, the viral load dynamic of Omicron infected individuals presented considerable age variation and clinical severity variation. For some specific workplaces, such as aging healthcare facilities and schools, the graded control measures should be recommended due to possible increased clinical severity. Initial Ct value could forecast severe outcome. Unvaccinated older people over 60 years of age with specific comorbidities are the high-risk groups associated with higher viral load and longer shedding duration. People with mild-moderate symptoms were less contagious after symptom resolution, but for severe or above patients, contagious characteristics could sustain for more than 1 week. To prevent worsened consequences, timely using of antiviral drugs within 3 days since onset was recommended targeting mild-moderate patients.

### Ethics approval

This study was performed in compliance with an institutional review board protocol at Huashan Hospital (KY2022-596). Written informed consent was obtained from each participant.

## Availability of data and materials

The data analyzed are not publicly available as they contain personal information.

## Authors’ contributions

Conception or design of the work: HJY, WHZ

Data collection: YL, JWA, SJG, HCZ, JPC, HYW, FS, YPW, JXZ, YW, RL, ZF, YLZ, XZ, XYYX, XMY, YCZ

Data analysis and interpretation: JXZ, JWA, YL, FS, SJG, HCZ Drafting the article: JXZ, JWA, YL

Critical revision of the article: HJY, WHZ, JXZ, JWA, YL, FS, SJG, HCZ, JJZ

Final approval of the version to be published: All of the authors.

## Data Availability

The data analyzed are not publicly available as they contain personal information.

## Acknowledgements

Hongjie Yu acknowledges financial support from the Key Program of the National Natural Science Foundation of China (82130093). Wenhong Zhang, Juanjuan Zhang acknowledge financial support from Shanghai Municipal Science and Technology Major Project (HS2021SHZX001). Wenhong Zhang acknowledges financial support from the Key Program of the National Natural Science Foundation of China (82161138018 and 82041010), Shanghai Science and Technology Committee (20dz2260100, 20Z11901100) and Shanghai Hospital Development Center (SHDC2020CR5010-002). We thank all physicians and patients for participating in this study. We appreciate all volunteers for helping collect data.

