## supplementary material for "Viral load dynamics in asymptomatic and symptomatic patients during Omicron BA.2 outbreak in Shanghai, China, 2022: a longitudinal cohort study"

### Nucleic Acid Sampling and Detection Procedure

Clinicians were required to insert the soft tip of the swab into the patient’s nostril and gently twist the swab against the inner wall of the nasal vestibule. The nasopharyngeal swabs were subsequently placed in a collection tube containing preserving fluid. The test procedure was as follows: purified viral nucleic acid (RNA) was extracted from the preserving fluid. Subsequently, an RT-PCR assay was performed after sampling, following the manufacturer’s instructions, including preparing the reaction system, adding the treated sample, and then amplification was performed on a real-time quantitative thermal cycler. Finally, the cycle threshold (Ct) values of both ORF1ab and N genes of SARS-CoV-2 were collected.

Patients discharged after two consecutive negative results(cycle threshold value of both ORF1ab and N gene >35).

### Additional results

#### Viral shedding dynamic by age and clinical severity

Further, we compared Ct value, viral shedding duration stratified by both clinical severity and age group (fig S2). For asymptomatic cases and mild-moderate patients, the older age groups had higher viral loads and longer viral shedding durations, while, the age effect was unobvious for severe and above patients (FigS2.A). Specifically, for asymptomatic cases, the average Ct values of 0-17 and 18-59 years old group were both more than 35 since initial detection with viral shedding durations around 5 days, while the average Ct value were less than 35 for patients more than 60 years old with VST around 6 days. For mild-moderate patients, the VST had correspondingly prolonged with 6.1 (IQR: 4.1-8.6), 6.3 (IQR: 4.4-8.8), 7.4 (IQR: 5.0-10.4) and 9.6 (IQR: 6.3-13.3), respectively. Additionally, for patients with positive Ct value, the times of peak Ct value accurance were similar among groups (4-5 days) but peak concentrations showed gradual increase with age and clinical severity (*p* <0.001) (FigS2.C).

### Supplement figures and tables

Table S1. Characteristics between participants and non-participants.

| Characteristics | Participants  (n=48600, 99.5%) | Excluded patients  (n=230, 0.5%) | *p value* |
| --- | --- | --- | --- |
| Clinical group |  |  |  |
| Asymptomatic | 15101 (31) | 64 (28) | <0.001 |
| Mild-moderate | 32909 (68) | 129 (56) |  |
| Severe | 461 (1) | 26 (11) |  |
| Critical or death | 129 (0) | 11 (5) |  |
| Gender |  |  |  |
| Female | 30518 (63) | 146 (63) | 0.884 |
| Male | 18082 (37) | 84 (37) |  |
| Age (years) |  |  |  |
| Median (IQR) | 45 (32-55) | 50 (37-65) |  |
| 0-17 | 1809 (4) | 2 (1) | 0.000 |
| 18-59 | 39680 (82) | 156 (68) |  |
| 60-79 | 6478 (13) | 58 (25) |  |
| 80+ | 633 (1) | 14 (6) |  |
| No. of comorbidities |  |  |  |
| None | 37951 (78) | 142 (62) | <0.001 |
| 1-2 | 7973 (16) | 48 (21) |  |
| ≥3 | 2103 (4) | 39 (17) |  |
| Missing | 573 (1) | 1 (0) |  |
| Vaccination type |  |  |  |
| Unvaccinated | 5443 (11) | 59 (26) | <0.001 |
| Inactivated_Partial (1 dose) | 1475 (3) | 8 (3) |  |
| Inactivated_Full (2 doses) | 16789 (35) | 59 (26) |  |
| Inactivated_booster (3 doses) | 22601 (47) | 93 (40) |  |
| Ad_Full (1 dose) | 937 (2) | 0 (0) |  |
| Ad_booster (2 doses) | 440 (1) | 2 (1) |  |
| Protein_Partial (<3 doses) | 33 (0) | 0 (0) |  |
| Protein_Full (3 doses) | 289 (1) | 3 (1) |  |
| Mixed_Partial/Full (2/3 doses) | 12 (0) | 0 (0) |  |
| Mixed_Booster (3 doses) | 13 (0) | 0 (0) |  |
| Missing | 568 (1) | 6 (3) |  |

^#^ Excluded patients due to extremely long virus shedding time (VST > 30 days).
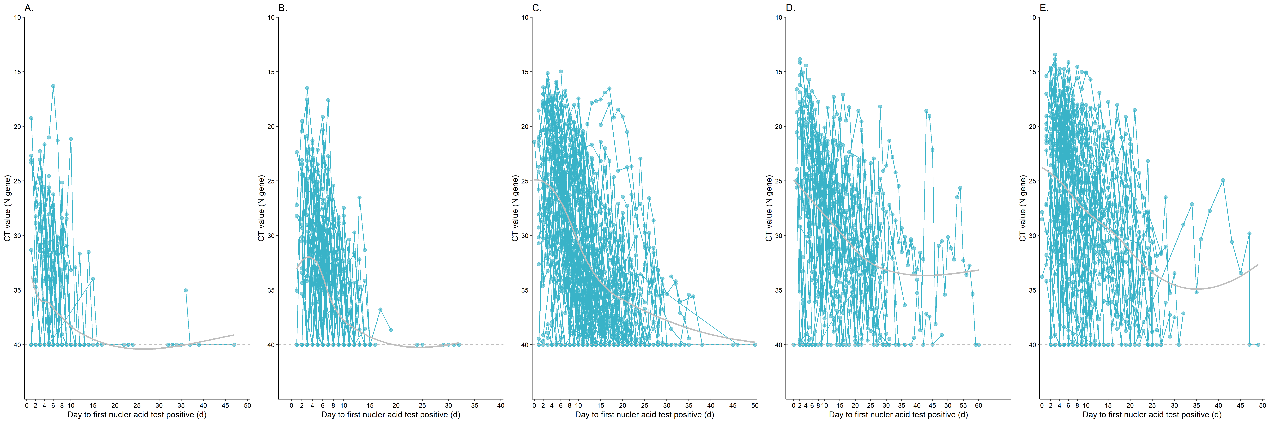

Fig S1. Individual profile of personal Ct value since 1st positive PCR test.

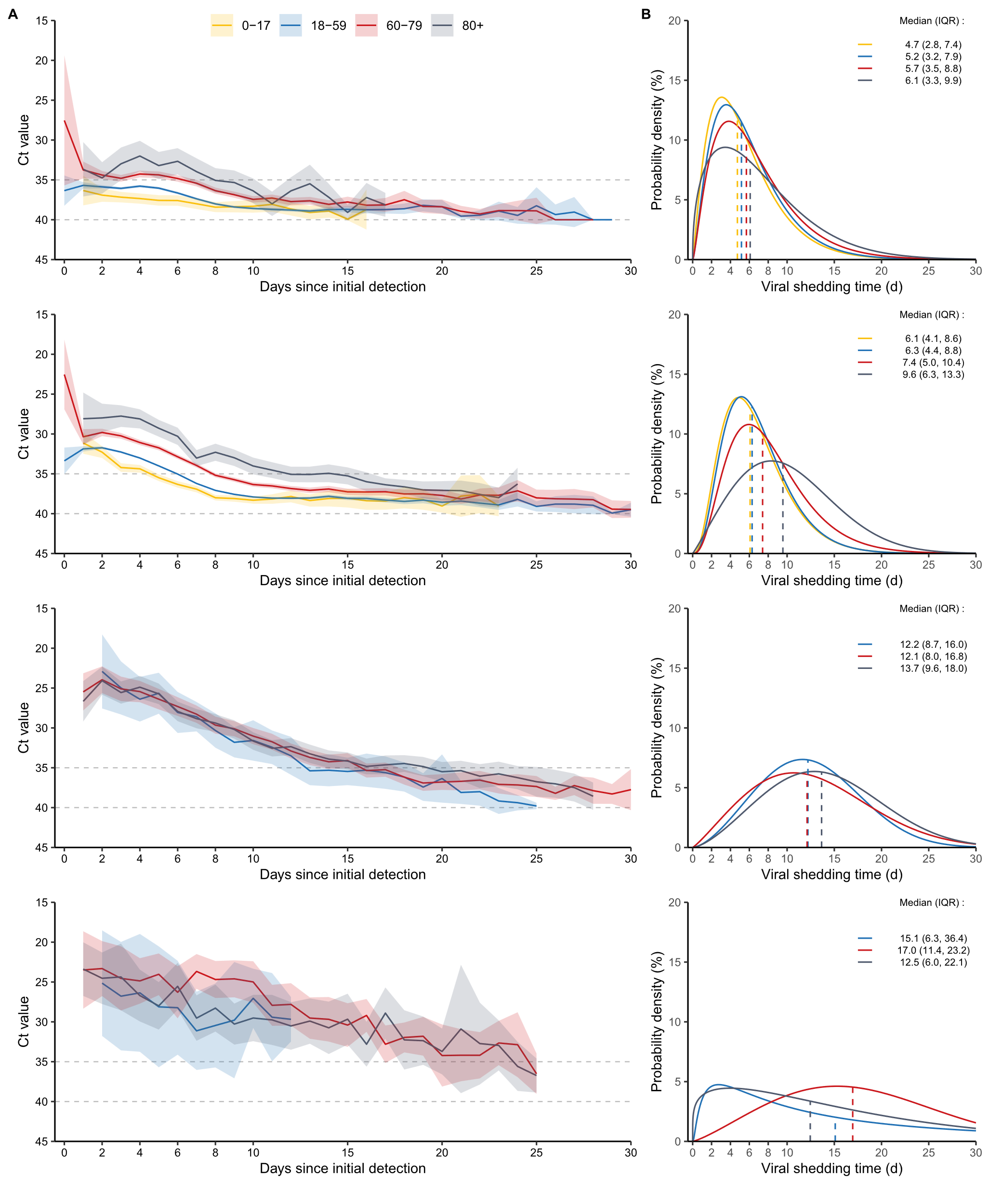

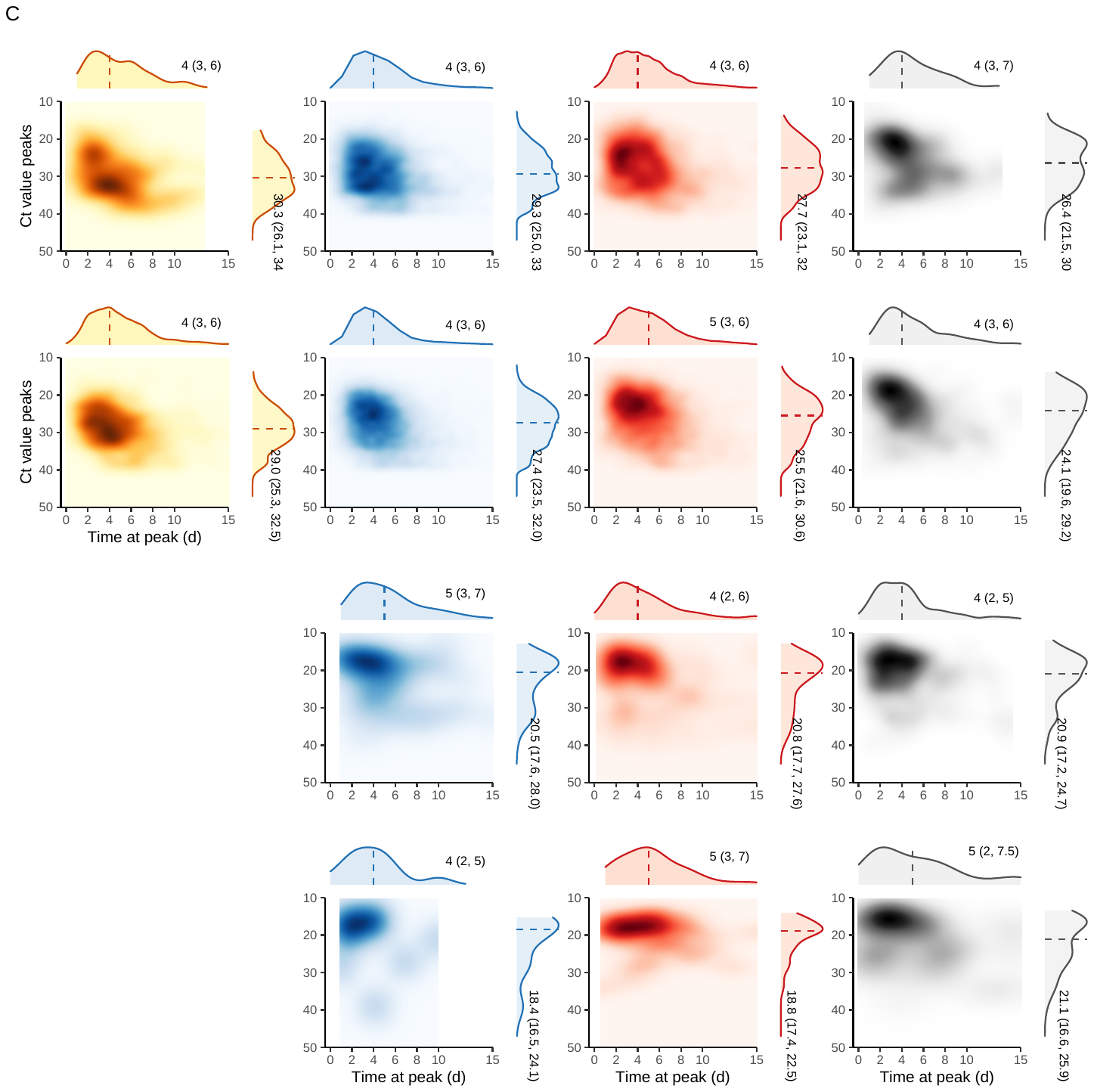

Fig S2. Clinical severity-specific and age group-specific characteristics of viral shedding. (A) Ct value at each time point since first positive PCR test. Solid line indicates average Ct value, shadow indicates 95% CI. (B) Time distribution of viral shedding duration. Vertical dashed line indicates median of viral shedding time. (C) Correlation between peak Ct value and the time to reach peak since first positive PCR test. Vertical dashed line indicates median of time at peak and the median of peak Ct value The footnote of alphabet indicates clinical severity groups including asymptomatic group, mild-moderate group, severe group, and critical or death group, respectively in sequence.

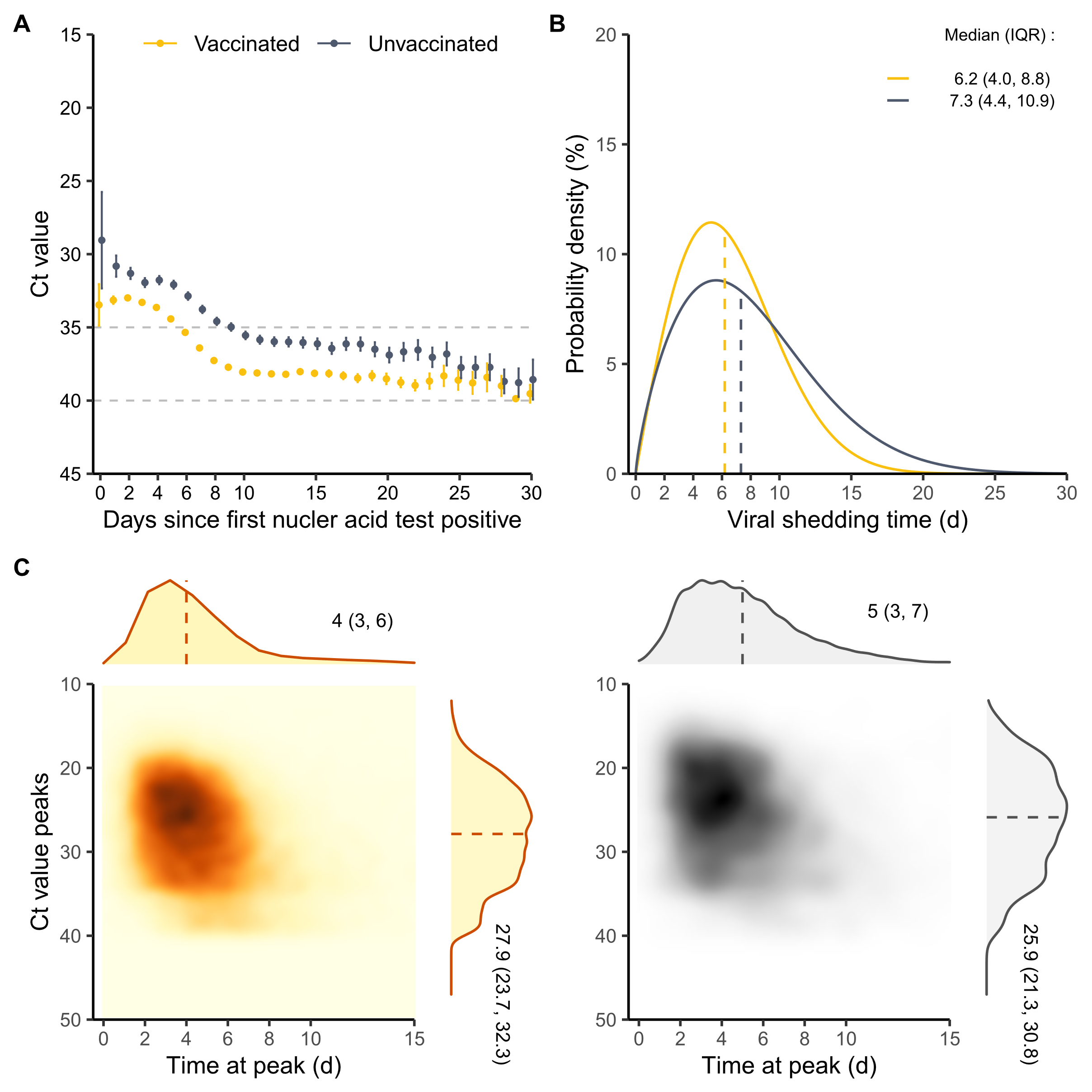

Fig S3. Vaccination status-specific characteristics of viral shedding. (A) Ct value at each time point since first positive PCR test. Vertical bar of each points means 95% confidence interval. (B) Time distribution of viral shedding duration. Vertical dashed line indicates median of viral shedding time. (C) Correlation between peak Ct value and the time to reach peak since first positive PCR test. Vertical dashed line indicates median of time at peak and the median of peak Ct value.

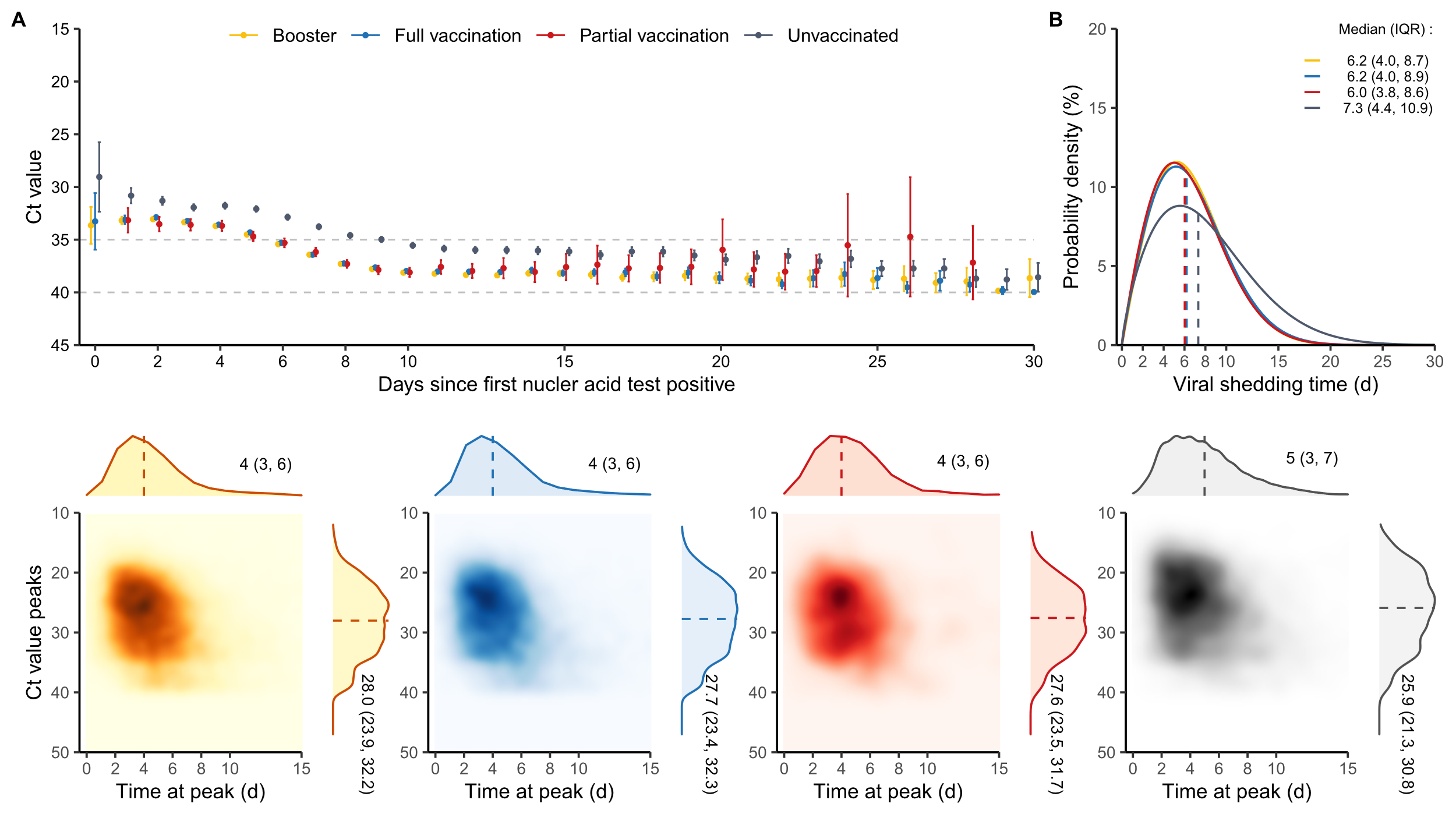

Fig S4. Vaccination status-specific characteristics of viral shedding of participants who vaccinated Ad vaccination or not. (A) Ct value at each time point since first positive PCR test. Vertical bar of each points means 95% confidence interval. (B) Time distribution of viral shedding duration. Vertical dashed line indicates median of viral shedding time. (C) Correlation between peak Ct value and the time to reach peak since first positive PCR test. Vertical dashed line indicates median of time at peak and the median of peak Ct value.

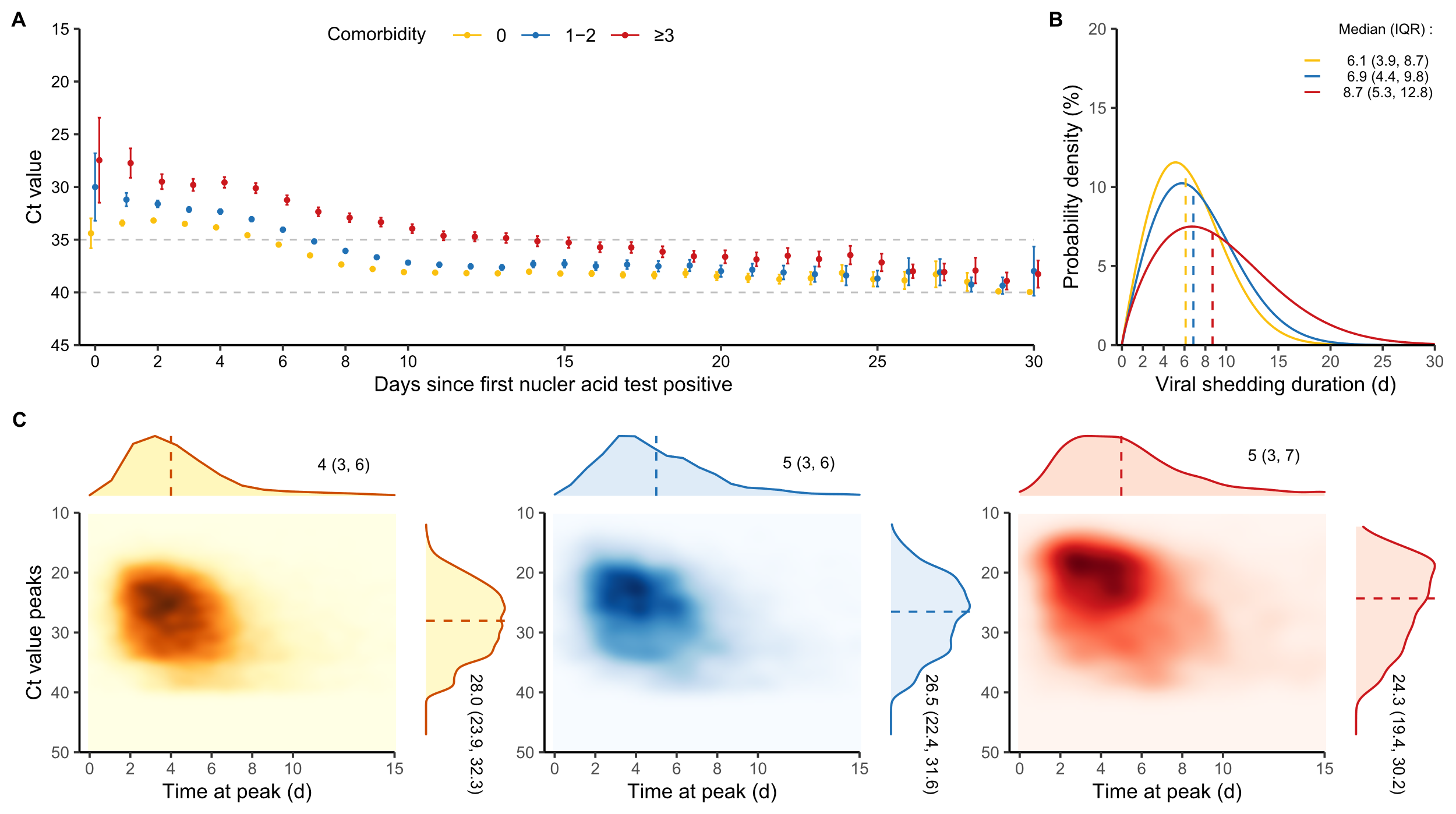
 Fig S5. Characteristics of viral shedding of participants with different number of comorbidities. (A) Ct value at each time point since first positive PCR test. Vertical bar of each points means 95% confidence interval. (B) Time distribution of viral shedding duration. Vertical dashed line indicates median of viral shedding time. (C) Correlation between peak Ct value and the time to reach peak since first positive PCR test. Vertical dashed line indicates median of time at peak and the median of peak Ct value.

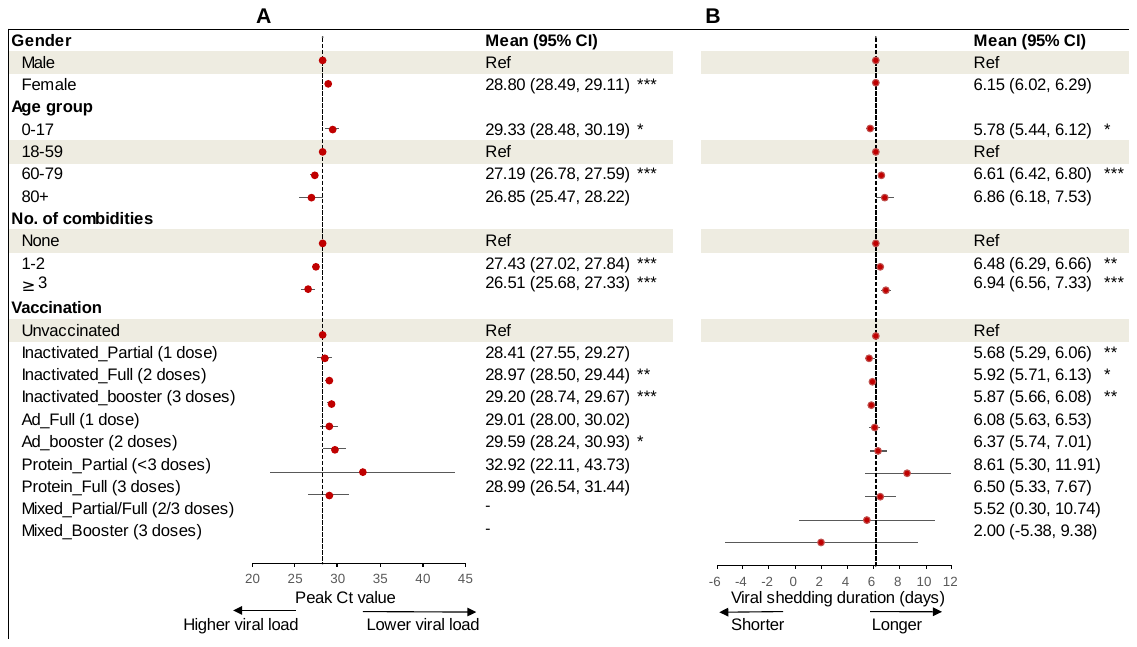
 Fig S6. Risk factors associated with peak viral load (A) and viral shedding duration (B) in asymptomatic cases. The coefficients (dots) and 95% confidence intervals (95% CIs, line segments) were estimated from a multivariable regression analysis with an adjustment for gender, age, number of comorbidities, and vaccination-infection status. Regression coefficients along with 95%CIs are reported as solid dots and horizontal lines relative to the value of the regression intercept. * p<0.05; ** p<0.01; *** p < 0.001.

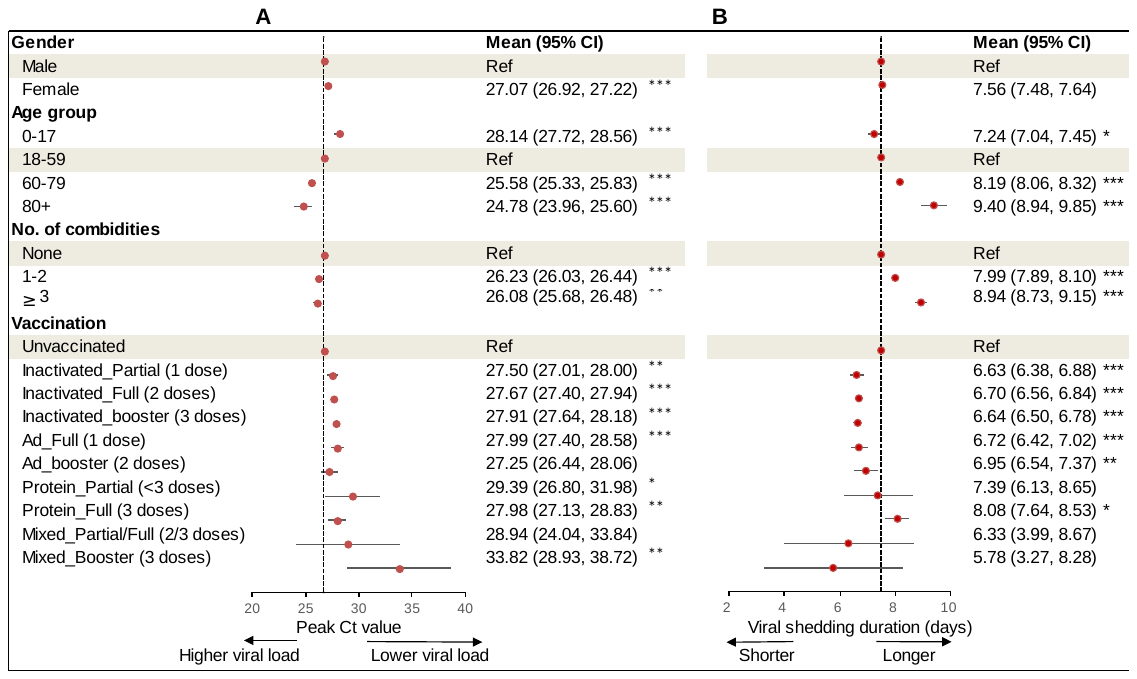

Fig S7. Risk factors associated with peak viral load (A) and viral shedding duration (B) in mild-moderate cases. The coefficients (dots) and 95% confidence intervals (95% CIs, line segments) were estimated from a multivariable regression analysis with an adjustment for gender, age, number of comorbidities, and vaccination-infection status. Regression coefficients along with 95%CIs are reported as solid dots and horizontal lines relative to the value of the regression intercept. * p<0.05; ** p<0.01; *** p < 0.001.

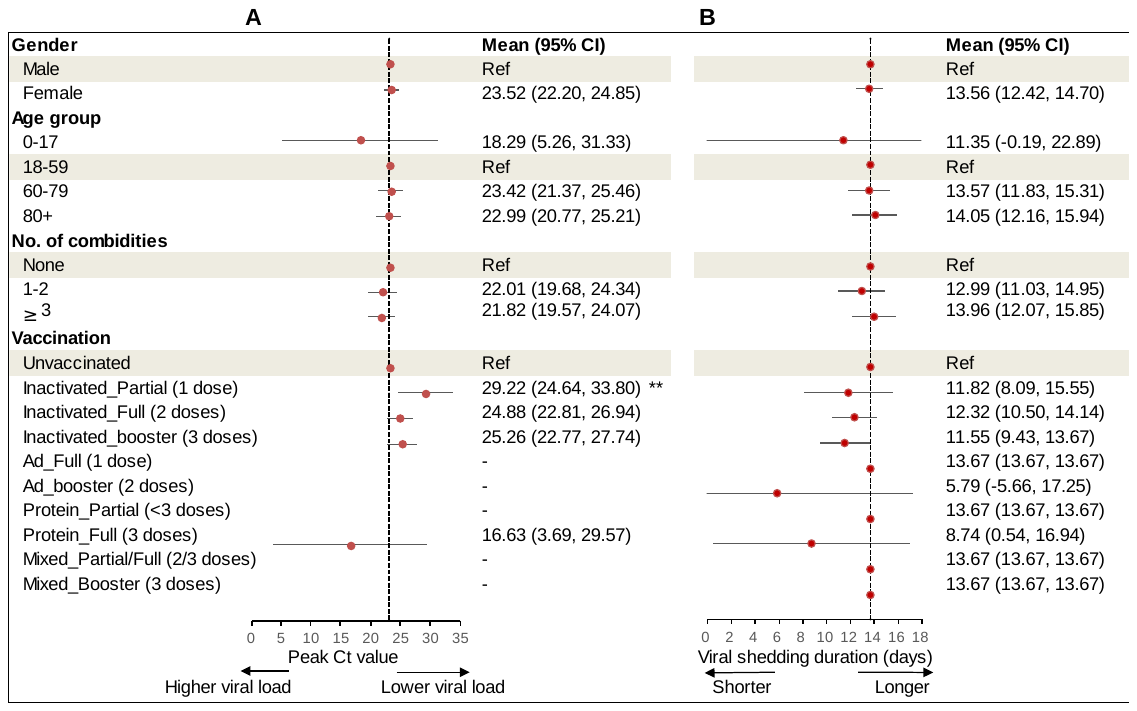

Fig S8. Risk factors associated with peak viral load (A) and viral shedding duration (B) in severe cases. The coefficients (dots) and 95% confidence intervals (95% CIs, line segments) were estimated from a multivariable regression analysis with an adjustment for gender, age, number of comorbidities, and vaccination-infection status. Regression coefficients along with 95%CIs are reported as solid dots and horizontal lines relative to the value of the regression intercept. * p<0.05; ** p<0.01; *** p < 0.001.
